# METACOGNITION, NETWORK-LINKED BEHAVIOURAL INDICES, AND PSYCHOSOCIAL FUNCTIONING: A MULTI-GROUP ESEM–SEM STUDY

**DOI:** 10.1101/2025.09.12.25335642

**Authors:** Vitalii Lunov, Mykhailo Matiash, Natalia Yevdokymova, Bohdan Tkach, Inha Rozhkova, Mykhailo Ilin, Andrii Pavlov, Volodymyr Prints

## Abstract

Metacognitive beliefs and behavioural indices aligned with large-scale networks plausibly shape day-to-day psychosocial functioning, especially under conditions of persistent stress and fatigue. However, their joint structure and group-level comparability remain under-specified. In a cross-sectional multi-group design (men and women), we administered the MCQ-30, the Fatigue Severity Scale, behavioural proxies of fronto-parietal, salience and default-mode network tendencies, and the Index of Psychosocial Functioning (IPF). Note that the three neurocognitive instruments do not measure large-scale brain networks per se. They are behavioural proxies that characterise probable patterns of attention/control, self-referential processing, and cue detection/switching putatively linked to activity within the frontoparietal (executive), default-mode, and salience networks; accordingly, all inferences are at the level of behaviour/cognition rather than direct neurophysiology. Using multi-group exploratory structural equation modelling (ESEM) with target rotation, we specified a three-factor measurement solution for the metacognitive– behavioural indicators, tested configural, metric and partial scalar invariance across sex, and regressed a latent IPF outcome on the three factors while controlling for age and education. Missingness was handled with FIML and MLR; WLSMV sensitivity analyses and bootstrap confidence intervals were used where appropriate. The three-factor ESEM structure showed good fit in each group and supported metric and partial scalar invariance. A factor reflecting threat/uncontrollability beliefs displayed robust negative associations with psychosocial functioning, whereas executive confidence/goal-directedness was positively associated; self-focus/monitoring contributed little or inconsistently. Effects were robust to estimator choice and to freeing a small subset of intercepts for partial scalar invariance. Clean separation of predictors and outcome (IPF as a latent criterion) clarifies that metacognitions centred on perceived threat and loss of control undermine functioning, while executive confidence aligned with FPN-like behaviour supports it. Therapeutically, the findings prioritise dampening uncontrollability beliefs and strengthening executive stamina. Behavioural network indices should be interpreted as proxies rather than neural measurements.

## Introduction

Chronic fatigue syndrome (CFS/ME) is characterised by persistent, disabling fatigue, disproportionate post-exertional symptom exacerbation, and a suite of cognitive and psychosocial difficulties that together erode everyday functioning. While argument continues over aetiology and optimal treatment, converging evidence indicates that the syndrome is sustained by interacting processes across levels of analysis: metacognitive beliefs that amplify symptom monitoring and emotional dysregulation; domain-specific inefficiencies in attention, processing speed, and episodic memory; and psychosocial disruptions that are often compounded by the “invisibility” of the illness in clinical and relational contexts (Cockshell & Mathias, 2010; Maher-Edwards et al., 2011, 2012; Pilkington et al., 2020). Rather than competing accounts, these strands appear mutually reinforcing. Theoretical and empirical work increasingly suggests that CFS/ME reflects disturbances of self-regulation spanning cognitive, emotional, social, and immunological systems - a perspective that demands integrative measurement and modelling (Capobianco et al., 2020; Lenzo et al., 2020; Raanes & Stiles, 2021; Walitt et al., 2024).

At the level of metacognition, patients commonly endorse negative beliefs regarding the uncontrollability and danger of thoughts, diminished cognitive confidence, and a perceived need to control mental content. These beliefs are associated with symptom severity and functional impairment, even when anxiety and depression are controlled, and they are theorised to maintain a cognitive-attentional syndrome of worry, rumination, and hypervigilant symptom monitoring (Maher-Edwards et al., 2011, 2012; Wells & Cartwright-Hatton, 2004). Recent transdiagnostic evidence in medical populations further links metacognitive constructs to emotional distress and reduced quality of life, suggesting that such beliefs are not epiphenomena but plausible drivers of disability (Capobianco et al., 2020; Lenzo et al., 2020). Longitudinal and multi-study analyses also indicate that beliefs about uncontrollability prospectively predict maladaptive regulation strategies and clinical symptoms, underscoring their mechanistic relevance (Salguero & Ramos-Cejudo, 2023; Strand et al., 2023).

Neurocognitive findings delineate a characteristic but heterogeneous profile. Meta-analytic and systematic reviews identify reliable impairments in information-processing speed, selective/divided attention, reading speed, and aspects of episodic memory, with instrumental functions often spared (Cockshell & Mathias, 2010; Aoun Sebaiti et al., 2022). Crucially, subjective cognitive complaints and objective performance are only modestly coupled, implying that metacognitive appraisals and perceived effort contribute importantly to the lived experience of “brain fog” (Rasouli et al., 2019). Beyond test scores, mechanistic accounts emphasise a shift from automatic to controlled processing modes under pain, fatigue, and interoceptive monitoring, with consequent slowing, distractibility, and an exaggerated sense of mental effort (Teodoro et al., 2018). Comparative work suggests that the cognitive phenotype of CFS/ME is not reducible to comorbid conditions; for instance, head-to-head data with multiple sclerosis point to both overlaps and a distinct consolidation deficit in CFS/ME (Aoun Sebaiti et al., 2025). Recent deep phenotyping of post-infectious CFS/ME also highlights alterations in effort preference and integrative brain systems, alongside immune signatures, drawing catecholaminergic and autonomic mechanisms into view (Walitt et al., 2024).

Psychosocial functioning is the arena in which these processes accrue salience for patients’ lives. Qualitative syntheses depict a relational illness marked by delegitimation, loss of identity, and strained encounters with services - dynamics that cannot be reduced to fatigue per se (Pilkington et al., 2020). Quantitative reviews identify associations between emotion regulation, executive functioning, sleep, and pro-inflammatory cytokines (e.g., IL-1, IL-6, TNF-α), though longitudinal evidence remains sparse (Raanes & Stiles, 2021). Together, these data motivate models in which cognition, metacognition, and immune activity are reciprocally coupled, shaping participation in work, education, relationships, and self-care. They also problematise unitary cognitive-behavioural accounts that attribute perpetuation chiefly to illness beliefs and avoidance, a critique that calls for theory and interventions proportionate to the biopsychosocial complexity of the syndrome (Geraghty et al., 2019), even while acknowledging that cognitive-behavioural packages can yield benefits for some patients and that outcomes vary with patient characteristics (Kuut et al., 2023).

Against this backdrop, there is a methodological gap. Many studies examine one domain in isolation, or collapse heterogeneous indicators into single composites. Fewer adopt an explicitly integrative psychometrics that align metacognitions, executive and salience-related functions, fatigue burden, and psychosocial role impairment within a single latent framework, and fewer still examine whether this framework differs by sex. Given the known heterogeneity of CFS/ME and the mixed literature on moderators of psychological outcomes, sex-stratified modelling is warranted to test whether the same instruments resolve different latent constellations across men and women. Doing so is clinically meaningful: if latent architectures diverge, then mechanistic targets - and thus treatment emphases - should be tailored accordingly.

The present study addresses this gap by combining validated measures of metacognitive beliefs, behavioural proxies of core neurocognitive networks, fatigue severity, and psychosocial functioning in a sizeable adult cohort with CFS/ME. Metacognitions were assessed with the MCQ-30, preserving its five subscales - (lack of) cognitive confidence, positive beliefs about worry, cognitive self-consciousness, negative beliefs about uncontrollability/danger, and need to control thoughts (Wells & Cartwright-Hatton, 2004). Executive/frontoparietal, default-mode, and salience-network functions were indexed with Ukrainian measures designed as behavioural proxies of these systems across defined domains, enabling fine-grained examination of sustained attention, working memory, problem solving, temporal integration, social-bond activation, and cue-driven switching (Sereda & Lunov, 2024, 2024a, 2024b). Fatigue was captured with the Fatigue Severity Scale, and psychosocial functioning across work, education, relationships, and self-care was measured using the Inventory of Psychosocial Functioning (IPF) framework, including a validated brief Ukrainian adaptation (Marx et al., 2020; Alexina et al., 2024). Our theoretical review anticipated specific couplings: metacognitive dyscontrol with reduced executive efficiency and memory clarity; salience-network switching with activity initiation and role maintenance; and global fatigue with both cognitive and role-function limits (Aoun Sebaiti et al., 2022; Maher-Edwards et al., 2011, 2012; Teodoro et al., 2018; Walitt et al., 2024).

### Study Aim

This study aimed to clarify how metacognitive beliefs and behavioural indices aligned with large-scale network tendencies cohere into a stable measurement structure and how these latent dimensions relate to psychosocial functioning. Specifically, we sought to (i) establish a three-factor ESEM solution for the metacognitive–behavioural indicators and test its multi-group invariance across men and women, and (ii) estimate the structural associations between these latent factors and a latent Index of Psychosocial Functioning (IPF) while cleanly separating predictors from the outcome and adjusting for age and education. A secondary aim was to examine whether fatigue partially mediates the association between threat/uncontrollability beliefs and functioning, and to verify robustness against estimator choice and a strict CFA alternative.

### Hypotheses

We hypothesised that a three-factor ESEM structure - capturing executive confidence/goal-directedness, threat/uncontrollability, and self-focus/monitoring - would fit well in each sex and achieve at least metric, and likely partial scalar, invariance across groups. We further hypothesised that executive confidence/goal-directedness would be positively associated with latent psychosocial functioning, whereas threat/uncontrollability would be negatively associated, with self-focus/monitoring showing little to no unique contribution once the other factors are controlled. Finally, we expected that fatigue would partially mediate the detrimental effect of threat/uncontrollability on functioning and that the magnitudes of structural paths would not materially differ between men and women.

### Metacognitive Beliefs in Chronic Fatigue Syndrome

The literature on chronic fatigue syndrome (CFS) has consistently underscored the role of cognitive appraisals in shaping illness perception and symptom burden. Yet, it is only within the last two decades that researchers have moved beyond first-order cognitions - such as beliefs about fatigue itself - to examine the influence of metacognitions, or beliefs about one’s cognitive processes. Metacognition, as defined in Wells’ Self-Regulatory Executive Function (S-REF) model (Wells, 2009), encompasses higher-order appraisals regarding the controllability, utility, or danger of cognitive activity. In the context of CFS, these metacognitions appear to be central not only to symptom severity but also to the broader psychosocial functioning of affected individuals.

Empirical evidence indicates that dysfunctional metacognitive beliefs are prominent in individuals with CFS. Maher-Edwards et al. (2011) demonstrated that negative metacognitions - particularly beliefs about the uncontrollability and dangerousness of thoughts - were stronger predictors of symptom severity than comorbid anxiety or depression. In this study, cognitive confidence and the perceived need to control thoughts predicted fatigue and functional impairment independently of emotional distress. This suggests that metacognitions are not secondary correlates of affective states but exert an independent influence on the clinical profile of CFS. A subsequent investigation by Maher-Edwards et al. (2012) extended this line of inquiry by constructing metacognitive profiles of CFS patients, revealing a pattern of both positive and negative metacognitions. Patients frequently endorsed beliefs that worrying could help them cope with symptoms, while simultaneously fearing that worry was uncontrollable and potentially harmful. This ambivalent configuration illustrates a paradox: metacognitive strategies may be initially adopted for adaptive purposes, yet their chronic activation engenders maladaptive emotional and functional consequences.

The broader metacognitive literature across chronic medical conditions supports the specificity of these findings. Capobianco et al. (2020), in a systematic review of physical illnesses, concluded that negative metacognitive beliefs concerning the uncontrollability and danger of thoughts are reliably associated with heightened anxiety and depression across diverse conditions. Similarly, Lenzo et al. (2020) reported that dysfunctional metacognitions predict reduced quality of life independent of illness severity in a range of chronic diseases. These transdiagnostic findings reinforce the relevance of metacognitive models for CFS, situating the syndrome within a broader framework of cognitive-attentional dysregulation observed in chronic health conditions.

Recent research has provided further nuance to these observations. Salguero and Ramos-Cejudo (2023), examining metacognitive beliefs about uncontrollability across multiple studies, confirmed that such beliefs uniquely predict maladaptive emotion regulation strategies and long-term symptom trajectories. Strand et al. (2023) similarly demonstrated that changes in metacognitive beliefs are stronger within-person predictors of depressive symptoms over time than shifts in dysfunctional attitudes. These findings not only affirm the predictive value of metacognition in psychopathology but also underscore its dynamic role in symptom fluctuation, lending weight to its clinical salience in conditions such as CFS.

The clinical implications of metacognitive dysfunction in CFS remain contentious. Traditional cognitive-behavioural therapy (CBT) approaches have attempted to modify illness beliefs and activity patterns, but their theoretical foundation - the so-called “cognitive behavioural model” of CFS - has been sharply criticised. Geraghty et al. (2019) argued that the model lacks empirical robustness, fails to account for accumulating biomedical evidence, and risks invalidating patient experiences by framing symptoms as perpetuated primarily by irrational beliefs. This critique highlights a tension between metacognitive conceptualisations and reductionist behavioural models. While CBT has demonstrated efficacy for some subgroups of CFS patients (Kuut et al., 2023), its benefits appear moderated by individual factors such as age, functional impairment, and self-efficacy. Such heterogeneity suggests that interventions targeting metacognition may require greater specificity than traditional CBT protocols afford.

Beyond CFS, studies of other neurological conditions further illuminate the clinical significance of metacognitive dysfunction. Heffer-Rahn and Fisher (2018), investigating multiple sclerosis, found that metacognitive beliefs accounted for emotional distress over and above demographic and clinical predictors, with negative beliefs about uncontrollability and danger making the largest contribution. These results suggest that metacognition exerts a transdiagnostic influence on adjustment to chronic illness, raising the possibility that targeted metacognitive therapy (MCT) could be a viable adjunctive approach in CFS.

The theoretical utility of focusing on metacognition lies in its capacity to bridge cognitive and emotional models of CFS. Rather than viewing fatigue solely as a biomedical phenomenon or a maladaptive behavioural response, the metacognitive perspective situates symptoms within the domain of higher-order self-regulation. Patients’ beliefs about their cognitive processes shape how they attend to bodily sensations, how they attempt to control symptoms, and how they interpret fluctuations in their condition. These processes, in turn, reinforce cycles of attentional bias, worry, and rumination that exacerbate perceived impairment. Metacognition thus provides a compelling explanatory construct for understanding the persistence of symptoms and for designing interventions that disrupt maladaptive cycles of self-monitoring and cognitive control.

### Neurocognitive Profiles and Cognitive Dysfunction in Chronic Fatigue Syndrome

Cognitive dysfunction is among the most disabling and frequently reported features of chronic fatigue syndrome (CFS), yet it remains one of the least well understood. Patients describe difficulties with attention, short-term memory, word-finding, and concentration, often referring to these experiences as “brain fog.” Importantly, such complaints are not epiphenomena of fatigue alone; a substantial body of neuropsychological research has identified objective impairments across specific cognitive domains that persist even after controlling for comorbid depression or pain. This growing literature suggests that CFS is associated with a distinct neurocognitive phenotype that carries profound implications for both diagnostic assessment and psychosocial functioning.

Meta-analytic evidence provides a robust synthesis of these findings. Cockshell and Mathias (2010), in one of the earliest meta-analyses of cognitive functioning in CFS, reported moderate to large deficits in information-processing speed, sustained attention, and working memory. By contrast, performance on measures of fine motor speed, vocabulary, and reasoning remained largely intact. These results pointed to a pattern of domain-specific rather than global impairment, challenging sceptical accounts that attributed cognitive complaints to non-specific factors such as low motivation or affective distress. Importantly, the deficits identified were consistent across multiple studies spanning two decades, underscoring their reliability.

More recent work has both confirmed and refined this cognitive profile. A systematic review and meta-analysis by Aoun Sebaiti et al. (2022) analysed nearly three decades of literature and highlighted impairments in visuospatial immediate memory, reading speed, and episodic verbal memory, particularly in retrieval and recognition processes. Attentional difficulties were also prominent, though executive functions appeared relatively preserved. This nuanced pattern suggests that CFS is characterised not by global cognitive decline but by targeted dysfunction in domains requiring rapid information processing and sustained attentional control. Such findings resonate with patient reports of cognitive effort, wherein even routine mental tasks are experienced as disproportionately exhausting.

The subjective perception of effort has become a key focus of contemporary accounts of CFS neurocognition. Teodoro, Edwards, and Isaacs (2018), in a systematic review of functional neurological disorders, fibromyalgia, and CFS, observed a consistent discordance between high rates of subjective cognitive symptoms and inconsistent objective deficits. They hypothesised that excessive interoceptive monitoring and a shift from automatic to effortful cognitive processing may underlie these experiences. Within this framework, patients’ complaints of “effortful cognition” are not simply exaggerated perceptions but may reflect a genuine neuropsychological mechanism: a maladaptive reallocation of attentional resources that impairs efficiency. This interpretation aligns with evidence from CFS cohorts indicating that cognitive performance is often inversely related to pain and fatigue, suggesting that subjective states modulate cognitive load in complex ways.

The relationship between subjective complaints and objective deficits remains a contested issue. Rasouli et al. (2019), in a large outpatient sample, found only weak correlations between self-reported cognitive difficulties and performance on neuropsychological tests. While nearly 40% of patients performed below clinical cut-offs on measures of attention and working memory, their subjective complaints were more strongly associated with fatigue, pain, and depression than with test scores. These results imply that subjective and objective cognitive impairment represent partially dissociable phenomena. From a clinical standpoint, this dissociation complicates diagnosis: reliance on subjective reports alone risks both over- and under-estimating impairment. Conversely, exclusive reliance on standardised tests may fail to capture the experiential reality of cognitive dysfunction in CFS.

Comparative studies have added further depth by situating CFS within a broader landscape of chronic illnesses. Aoun Sebaiti et al. (2025) compared cognitive profiles of patients with CFS and multiple sclerosis (MS). Both groups demonstrated deficits in episodic memory retrieval, visual selective attention, and reading speed, but CFS patients showed distinct impairments in consolidation processes. Crucially, these deficits did not correlate with fatigue, pain, or depression in either group, reinforcing the view that CFS entails intrinsic cognitive dysfunction. Such findings help to differentiate CFS from other neuroinflammatory conditions and may aid in refining diagnostic criteria, particularly in cases where fatigue alone does not provide adequate specificity.

Historical perspectives underscore both the persistence and evolution of this research trajectory. Early reviews, such as DiPino and Kane (1996), already documented attention and memory deficits in CFS, though methodological inconsistencies limited generalisability. Qualitative investigations from the same period revealed patients’ strong conviction that their illness was primarily physical, with cognitive complaints perceived as secondary to an underlying somatic pathology (Clements et al., 1997). These early tensions between subjective experience and clinical assessment continue to shape contemporary debates about the ontological status of cognitive dysfunction in CFS.

Contemporary research has shifted towards more sophisticated phenotyping approaches. Walitt et al. (2024), in a large-scale deep phenotyping study of post-infectious CFS, identified alterations in effort preference rather than central fatigue per se. Their findings implicated dysfunction in integrative brain regions associated with catecholaminergic pathways and autonomic regulation. Such abnormalities may explain why patients experience disproportionate cognitive effort and difficulty sustaining attention. Importantly, these neurobiological insights are accompanied by evidence of immune dysregulation, reinforcing the view that cognitive dysfunction in CFS arises from complex psychoneuroimmunological interactions.

The theoretical implications of this body of work are significant. First, CFS cannot be reduced to subjective fatigue alone: cognitive deficits are objectively demonstrable and domain-specific. Second, the dissociation between subjective complaints and objective performance underscores the need for multimodal assessment, incorporating both neuropsychological testing and patient-reported outcomes. Third, the distinctiveness of the CFS cognitive profile, relative to conditions such as MS, supports the conceptualisation of CFS as a disorder with its own neurocognitive signature. Finally, emerging neurobiological evidence suggests that abnormalities in effort regulation and interoceptive monitoring may underlie the subjective experience of “brain fog,” offering a plausible mechanistic bridge between cognition, symptom perception, and immune dysfunction.

In sum, the neurocognitive literature on CFS has matured from early descriptive accounts to a more refined understanding of domain-specific impairments, dissociations between subjective and objective findings, and putative neurobiological underpinnings. These advances provide a critical foundation for integrative models that link metacognitive beliefs, cognitive dysfunction, and psychosocial impairment - a task to which the following section now turns.

### Psychosocial Functioning and Psychoneuroimmunological Links

Beyond fatigue and cognitive dysfunction, chronic fatigue syndrome (CFS) exerts a profound toll on psychosocial functioning. Patients commonly report difficulties in sustaining interpersonal relationships, maintaining employment, and preserving a coherent sense of identity. These disruptions are not merely secondary consequences of prolonged fatigue but are themselves constitutive features of the syndrome, shaping both its clinical expression and its lived experience. Contemporary research indicates that psychosocial dysfunction in CFS is intertwined with metacognitive and neurocognitive factors, and increasingly, with immunological processes that influence both emotional regulation and social adaptation.

Qualitative research has illuminated the relational and identity-based challenges faced by individuals with CFS. Pilkington et al. (2020), in a meta-ethnography of patient narratives, described the syndrome as an “invisible illness” that undermines social recognition and corrodes the sense of self. Patients often experience alienation in healthcare encounters, frustration at the lack of visible biomarkers, and a sense of diminished legitimacy in the eyes of others. The result is a precarious negotiation of social roles, in which fatigue and cognitive difficulties disrupt employment, parenting, and intimate relationships. This relational paradigm underscores that psychosocial dysfunction in CFS cannot be understood solely as a downstream effect of somatic symptoms; it is embedded within broader cultural and interpersonal contexts that shape the illness trajectory.

Empirical findings reinforce this qualitative perspective. Deficits in emotion regulation, executive functioning, and sleep quality are consistently associated with diminished psychosocial adaptation. Raanes and Stiles (2021), in their systematic review, found that poor emotion regulation in CFS was linked to elevated levels of interleukin (IL)-2, while interpersonal difficulties were associated with IL-6 and tumour necrosis factor-alpha (TNF-α). Sleep disturbance, a near-universal complaint among patients, was correlated with multiple cytokines, including IL-1 and IL-6. These associations suggest that psychosocial functioning is not only a psychological construct but is biologically embedded through psychoneuroimmunological pathways. Dysregulated immune activity, manifesting in pro-inflammatory cytokine elevation, may amplify difficulties in emotional control, sleep architecture, and social interaction, thereby exacerbating psychosocial disability.

The convergence of psychological and immunological findings lends support to integrative models of CFS, yet the evidence remains methodologically constrained. As Raanes and Stiles (2021) emphasised, most studies are cross-sectional, precluding causal inference. Longitudinal designs are scarce, and sample sizes are often small. Nevertheless, the emerging picture is one in which psychosocial dysfunction, cognitive impairment, and immune dysregulation reinforce one another in a vicious cycle. Emotion dysregulation may amplify immune activation, which in turn worsens fatigue and cognitive symptoms, thereby deepening social withdrawal and identity disruption. Such circular causality highlights the limitations of reductionist models that privilege either psychological or biomedical explanations in isolation.

This tension is particularly evident in debates around the cognitive-behavioural model of CFS. The model posits that maladaptive illness beliefs and avoidance behaviours perpetuate fatigue and functional impairment, thereby justifying the use of cognitive-behavioural therapy (CBT) as a primary treatment. Yet, as Geraghty et al. (2019) argued, this framework risks invalidating patients’ experiences by framing psychosocial dysfunction as primarily the product of irrational cognitions. Moreover, it neglects accumulating evidence of immune and neurocognitive abnormalities that are not reducible to psychosocial processes. While CBT and related approaches can yield benefits for some patients (Kuut et al., 2023), the heterogeneity of outcomes underscores the inadequacy of any model that fails to account for the complex biopsychosocial ecology of CFS.

Emerging biological studies provide further depth to this integrative picture. Walitt et al. (2024), in their deep phenotyping of post-infectious CFS, identified abnormalities in effort preference and catecholaminergic pathways, alongside immune markers consistent with chronic antigenic stimulation. These findings suggest that psychosocial dysfunction is mediated not only by patients’ metacognitions or cognitive deficits but also by alterations in brain-immune communication. Dysfunction in integrative neural regions may impair the capacity to balance cognitive load and social demands, producing both subjective distress and objective functional impairment. This resonates with patients’ frequent descriptions of disproportionate effort in daily tasks, both cognitive and interpersonal.

The psychosocial impact of CFS is thus multifaceted. At the micro-level, difficulties in emotion regulation and executive functioning compromise day-to-day adaptation. At the meso-level, disrupted relationships and strained employment trajectories undermine social integration. At the macro-level, stigma and delegitimation reinforce isolation and psychological distress. Immunological dysregulation weaves through each of these levels, providing a biological substrate for psychosocial impairment. Importantly, metacognitive beliefs intersect with these processes: patients who believe their thoughts are uncontrollable or dangerous may engage in maladaptive monitoring of symptoms, amplifying both emotional distress and social withdrawal. In this way, metacognition acts as a mediator between neurocognitive dysfunction, psychosocial adaptation, and immune dysregulation.

Theoretical implications of this perspective are significant. Psychosocial dysfunction in CFS cannot be neatly categorised as a psychological consequence of fatigue, nor can it be reduced to immunological biomarkers. Instead, it reflects a dynamic interaction in which immune activation influences cognition and emotion, cognitive deficits constrain social functioning, and social adversity feeds back into metacognitive and immunological processes. This recursive interplay calls for models that transcend the binary of “psychogenic” versus “biomedical.” Instead, CFS should be conceptualised as a disorder of self-regulation across multiple systems - cognitive, emotional, social, and immunological.

From a clinical standpoint, this integrative framework suggests several directions. First, interventions should address not only symptom management but also the relational and identity-based dimensions of CFS, acknowledging the “invisible” nature of the illness. Second, psychotherapeutic approaches may benefit from incorporating metacognitive therapy, which targets beliefs about cognitive processes, rather than relying solely on behavioural modification. Third, collaboration between psychologists, neurologists, and immunologists is essential to develop multimodal interventions that address both psychosocial and biological determinants of disability. Only through such interdisciplinary integration can the complexity of psychosocial functioning in CFS be adequately understood and treated.

### Toward an Integrative Clinical and Theoretical Model

The preceding sections have traced three major strands of research in chronic fatigue syndrome (CFS): the centrality of dysfunctional metacognitive beliefs, the specificity of neurocognitive impairments, and the interplay of psychosocial and immunological variables. Each domain yields valuable insights into the phenomenology of the disorder. Yet, taken in isolation, none is sufficient to account for the complexity and chronicity of CFS. To advance theory and clinical practice, an integrative model is required - one that articulates how these domains intersect, mutually reinforce, and collectively sustain the syndrome.

At the metacognitive level, patients with CFS frequently endorse beliefs about the uncontrollability and danger of their thoughts, diminished cognitive confidence, and the necessity of controlling mental processes (Maher-Edwards et al., 2011, 2012; Capobianco et al., 2020). These beliefs foster maladaptive self-monitoring, worry, and rumination, which in turn intensify symptom perception. Crucially, they act as higher-order regulators of attentional deployment: patients preoccupied with their symptoms may allocate excessive cognitive resources to monitoring fatigue or cognitive lapses, thereby amplifying the subjective sense of effort. Such processes are not reducible to depression or anxiety; rather, they exert an independent influence on the severity and persistence of CFS symptoms (Strand et al., 2023; Salguero & Ramos-Cejudo, 2023).

The neurocognitive literature provides a mechanistic substrate for these metacognitive processes. Objective impairments in information-processing speed, attention, and episodic memory (Cockshell & Mathias, 2010; Aoun Sebaiti et al., 2022) constrain the efficiency with which patients can meet everyday cognitive demands. This inefficiency aligns with findings that cognitive tasks in CFS are experienced as disproportionately effortful (Teodoro et al., 2018; Walitt et al., 2024). The dissociation between subjective and objective impairments (Rasouli et al., 2019) further underscores the mediating role of metacognition: patients’ interpretations of their cognitive performance may exacerbate distress independently of measurable deficits. Moreover, comparative studies with multiple sclerosis suggest that the cognitive phenotype of CFS is distinct, particularly in relation to memory consolidation (Aoun Sebaiti et al., 2025), supporting the conceptualisation of CFS as a disorder with unique neurocognitive characteristics.

Psychosocial functioning constitutes the arena in which these cognitive and metacognitive processes manifest most visibly. Emotion regulation difficulties, interpersonal strain, and social isolation are hallmarks of the condition (Pilkington et al., 2020). Dysfunctional metacognitions magnify these difficulties by reinforcing cycles of symptom monitoring and withdrawal, while cognitive deficits reduce patients’ capacity to meet social and occupational demands. The resulting psychosocial disruption feeds back into cognitive processes, intensifying worry, hopelessness, and negative self-appraisals. Here, the social environment plays a crucial role: the invisibility of the illness and the delegitimation of patients’ experiences exacerbate distress, compounding the internal burden imposed by cognitive and metacognitive dysfunction.

Overlaying these domains is the immunological dimension. Dysregulated cytokine activity - particularly elevations in IL-1, IL-6, and TNF-α - has been associated with poor emotion regulation, impaired executive function, and disrupted sleep (Raanes & Stiles, 2021). Walitt et al. (2024) further implicated catecholaminergic pathways and B-cell abnormalities in post-infectious CFS, linking immune dysfunction to alterations in effort regulation and cognitive load. These findings suggest that immune activity both shapes and is shaped by psychological processes, providing a biological pathway through which psychosocial stressors may exacerbate symptoms. Indeed, chronic worry and emotional dysregulation may maintain low-grade inflammation, which in turn worsens fatigue and cognitive impairment, creating a self-reinforcing psychoneuroimmunological loop.

This integrative model thus envisions CFS as a disorder of self-regulation across multiple, interacting systems. At the cognitive level, patients exhibit specific neurocognitive impairments that limit efficiency. At the metacognitive level, they hold beliefs that amplify symptom monitoring and distress. At the psychosocial level, relational and identity-based disruptions erode resilience. At the immunological level, dysregulated cytokine activity exacerbates both cognitive and emotional dysfunction. Importantly, these domains do not operate in linear sequence but in recursive cycles: metacognitive worry sustains immune activation; immune dysregulation intensifies cognitive inefficiency; cognitive deficits fuel psychosocial withdrawal; and social isolation reinforces negative metacognitions.

The clinical implications of this model are far-reaching. First, it cautions against reductionist treatments that target only one domain - be it CBT for illness beliefs, pharmacological interventions for fatigue, or immune-modulating therapies. Instead, it advocates for multimodal interventions that address the full biopsychosocial ecology of the disorder. Metacognitive therapy may help patients disengage from maladaptive symptom monitoring; neurocognitive rehabilitation may strengthen attention and processing speed; psychosocial interventions may reduce stigma and support identity reconstruction; and psychoneuroimmunological approaches may modulate inflammatory activity. Second, the model underscores the need for personalised medicine: as Kuut et al. (2023) demonstrated, patient characteristics moderate treatment outcomes, suggesting that interventions must be tailored to metacognitive profiles, cognitive deficits, and immune markers.

Theoretically, the model advances our understanding of CFS by situating it at the intersection of cognitive science, clinical psychology, and psychoneuroimmunology. It bridges the gap between patients’ subjective experiences of fatigue and “brain fog,” objective findings of neurocognitive deficits, and emerging biomarkers of immune dysfunction. It also reframes the debate between psychogenic and biomedical accounts, proposing instead a systems-level perspective in which psychological and biological processes are mutually constitutive. In this sense, CFS exemplifies the need for post-reductionist models of illness, where mind and body are understood as components of dynamic regulatory networks.

Finally, the model has implications beyond CFS. The overlaps with fibromyalgia, functional neurological disorders, and post-viral syndromes such as long COVID (Teodoro et al., 2018; Walitt et al., 2024) suggest that similar self-regulatory dysfunctions may underpin a wider class of chronic, medically unexplained syndromes. By articulating how metacognition, cognition, psychosocial functioning, and immune processes interact, the integrative framework developed here may offer a template for understanding and treating these related conditions.

In sum, an integrative clinical and theoretical model of CFS must embrace the complexity of the disorder. It must acknowledge the central role of dysfunctional metacognitions, delineate the specificity of neurocognitive impairments, situate psychosocial dysfunction within relational and cultural contexts, and incorporate the emerging evidence of immune dysregulation. Only by weaving these domains into a coherent framework can researchers and clinicians move beyond piecemeal explanations towards a comprehensive understanding of CFS - one that honours patients’ experiences while advancing scientific knowledge and therapeutic innovation.

The theoretical review of chronic fatigue syndrome (CFS) presented here underscores the necessity of adopting an integrative perspective that transcends traditional dichotomies between “psychological” and “biological” explanations. The evidence demonstrates that dysfunctional metacognitive beliefs, domain-specific neurocognitive impairments, psychosocial disruption, and immune dysregulation each constitute essential components of the syndrome. Yet, these components do not operate in isolation. Rather, they form a network of mutually reinforcing processes that sustain symptomatology and functional impairment over time.

At the metacognitive level, patients with CFS frequently endorse beliefs about the uncontrollability and danger of their thoughts, diminished cognitive confidence, and the necessity of constant mental control (Maher-Edwards et al., 2011, 2012; Capobianco et al., 2020; Lenzo et al., 2020). These beliefs exacerbate symptom perception and reinforce maladaptive attentional strategies such as worry, rumination, and hypervigilant monitoring of fatigue. Importantly, metacognitive factors predict symptom severity independently of anxiety and depression (Strand et al., 2023; Salguero & Ramos-Cejudo, 2023), highlighting their centrality to the CFS phenotype.

The neurocognitive literature further reveals that impairments in processing speed, selective attention, and episodic memory are robust and reproducible (Cockshell & Mathias, 2010; Aoun Sebaiti et al., 2022). Patients frequently describe these deficits as “brain fog,” reflecting not only performance decrements but also an altered subjective experience of cognitive effort (Teodoro et al., 2018). Importantly, these impairments often show weak correlations with fatigue, pain, or depression (Rasouli et al., 2019; Aoun Sebaiti et al., 2025), reinforcing their status as autonomous clinical features. Comparative studies with multiple sclerosis and other chronic illnesses confirm that CFS carries a distinct neurocognitive signature.

Psychosocial functioning represents the domain in which these cognitive and metacognitive disturbances are most acutely felt. Relational strain, occupational impairment, and identity disruption are consistently reported by patients (Pilkington et al., 2020). These difficulties are intensified by stigma and delegitimation, which further erode resilience. At the same time, immune dysregulation - particularly elevations in IL-1, IL-6, and TNF-α - has been linked to poor emotion regulation, sleep disturbance, and interpersonal difficulties (Raanes & Stiles, 2021). Such findings demonstrate that psychosocial dysfunction in CFS is not a mere by-product of fatigue but is biologically embedded through psychoneuroimmunological pathways.

Synthesising these strands, the emerging picture is that of a recursive system: dysfunctional metacognitions amplify cognitive inefficiency; neurocognitive deficits constrain psychosocial functioning; psychosocial adversity heightens emotional distress; and immune activation reinforces cognitive and affective disturbances. This cyclical model challenges reductionist accounts and compels us to conceptualise CFS as a disorder of self-regulation across cognitive, emotional, social, and immunological systems. It also resonates with findings in related conditions such as fibromyalgia, functional neurological disorders, and long COVID, suggesting that a broader class of post-infectious and chronic syndromes may share similar regulatory disturbances (Teodoro et al., 2018; Walitt et al., 2024).

For clinical practice, these insights highlight the importance of multimodal interventions. Psychological therapies that target metacognitive processes, such as metacognitive therapy, may help patients disengage from maladaptive symptom monitoring. Neurocognitive rehabilitation could address attentional and memory deficits, while psychosocial interventions must confront stigma and support identity reconstruction. Finally, collaboration with immunology is essential, as immune-modulating approaches may ameliorate the biological underpinnings of fatigue and cognitive impairment. Crucially, treatment must be personalised: as meta-analytic evidence suggests, patient characteristics moderate response to interventions (Kuut et al., 2023).

The theoretical contribution of this review lies in articulating a model that neither dismisses the biological abnormalities observed in CFS nor reduces the illness to psychosocial constructs. Instead, it positions CFS as a paradigmatic example of a multi-system disorder in which mind and body are inseparable. Such a perspective is not merely academic; it has profound implications for diagnosis, prognosis, and the design of effective interventions. By embracing the complexity of CFS, clinicians and researchers can move towards a more comprehensive understanding that honours patient experiences while advancing scientific rigour.

In conclusion, chronic fatigue syndrome emerges from the interplay of metacognitive beliefs, neurocognitive inefficiencies, psychosocial adversity, and immune dysregulation. Each of these domains offers partial insight, but only when considered together do they reveal the self-perpetuating cycles that sustain illness. The challenge for future research is to translate this integrative framework into longitudinal, multimodal studies that can disentangle causal pathways and guide intervention. The challenge for clinical practice is to design treatments that are as multidimensional as the syndrome itself. In meeting these challenges, we may move closer to alleviating the profound burden of CFS for patients, families, and societies alike.

## Materials and Methods

Participants were recruited via community and academic mailing lists and completed an online battery. The final sample comprised N = 620 adults (men n = 300, women n = 320), aged M = 36.8, SD = 10.9 years. Inclusion criteria were age 18–65 and fluency in the study language; exclusion criteria were self-reported neurological disease or acute psychiatric crisis. Ethical approval was granted by the institutional review board, and all participants provided informed consent.

The battery included the Metacognitions Questionnaire-30 (MCQ-30; five 6-item subscales: Positive Beliefs about Worry, Negative Beliefs about Uncontrollability/Danger, Cognitive Confidence, Need to Control Thoughts, Cognitive Self-Consciousness), scored according to the manual. Fatigue was indexed by the nine-item Fatigue Severity Scale (FSS). Behavioural proxies of large-scale networks reflected fronto-parietal (FPN), salience (SN) and default-mode (DMN) tendencies, operationalised as brief composite indices of goal maintenance and distraction resistance (FPN), salience switching and interruptibility (SN), and self-referential focus and mind-wandering (DMN). These are behavioural proxies, not neuroimaging measures. Psychosocial functioning was measured with the Index of Psychosocial Functioning (IPF), comprising Work/Study, Self-Care, Family/Intimate and Friendships/Social domains. In structural models, IPF was specified as a latent outcome defined by these four domains, ensuring predictors and criterion were non-overlapping.

Analyses proceeded in a multi-group framework (men versus women). First, we estimated a three-factor ESEM for the metacognitive–behavioural indicators with target rotation that permitted theoretically small cross-loadings. We then tested configural, metric and partial scalar invariance across sex using conventional change thresholds (ΔCFI ≤ .010; ΔRMSEA ≤ .015). Secondly, the latent IPF outcome was regressed on the three factors, controlling for age and education; exploratory mediation via FSS used bias-corrected bootstrap confidence intervals with 5,000 resamples. The primary estimator was MLR with full-information maximum likelihood for missing data; sensitivity analyses using WLSMV for ordinal distributions and a strict CFA alternative (zero cross-loadings) assessed robustness. Reliability was indexed by McDonald’s ω; descriptive statistics and zero-order associations appear in Table 1. All syntax and decision logs are available on request.

**Table 1.** Descriptive statistics and reliability by sex.

| Scale / Domain | Men: M (SD) | Women: M (SD) | $\omega$ |
| --- | --- | --- | --- |
| MCQ – Positive Beliefs about Worry | 2.20 (0.58) | 2.26 (0.57) | .84 / .85 |
| MCQ – Negative Beliefs (Uncontrollability/Danger) | 2.78 (0.65) | 2.84 (0.66) | .88 / .89 |
| MCQ – Cognitive Confidence | 2.92 (0.56) | 2.86 (0.57) | .86 / .85 |
| MCQ – Need to Control Thoughts | 2.61 (0.62) | 2.68 (0.63) | .85 / .85 |
| MCQ – Cognitive Self-Consciousness | 3.01 (0.52) | 3.06 (0.51) | .82 / .83 |
| FPN proxy (goal maintenance) | 10.9 (3.0) | 10.6 (3.0) | .80 / .80 |
| SN proxy (salience switching) | 8.8 (3.1) | 9.1 (3.2) | .79 / .79 |
| DMN proxy (self-referential focus) | 9.3 (3.0) | 9.6 (3.1) | .78 / .79 |
| Fatigue Severity Scale (FSS) | 4.56 (1.28) | 4.71 (1.24) | .90 / .90 |
| IPF – Work/Study | 72.3 (14.5) | 70.9 (14.9) | - |
| IPF – Self-Care | 78.0 (12.7) | 77.2 (12.9) | - |
| IPF – Family/Intimate | 74.5 (14.8) | 73.8 (15.3) | - |
| IPF – Friendships/Social | 68.2 (15.7) | 66.9 (16.2) | - |

Note. MCQ subscales are reported as mean item scores (1–4). Higher FSS indicates greater fatigue. Higher IPF scores indicate greater psychosocial impairment. $\omega$ = McDonald's omega; men/women values shown separated by a slash.

### Measures (Domains/Subscales and Scoring)

Frontoparietal/Executive Network (FPN/CEN): Scale of Cognitive Assessment of the Executive Network (Sereda & Lunov, 2024). This instrument indexes putative executive network efficiency across four domains (five items each; 0–3): sustained attention; complex problem solving; working memory; and goal-directed behaviour and decision-making. Domain ranges are 0–15; the FPN total (Executive Network Index) is 0–60 with interpretive bands 0–20 (low), 21–40 (medium), and 41–60 (high).

Default Mode Network (DMN): Scale of Assessment of Passive Neurocognitive Activity (Sereda & Lunov, 2024a). This scale operationalises internally oriented functions attributed to the DMN across seven factors (five items each; 0–3): absorption of distractors; cognitive flexibility; depth of connection with self and world; activation of social-bond chains; temporal integration of past– present–future; creative self-expression; and clarification of vague memories. Factor sums are 0–15; the DMN total is 0–105 with interpretive bands 0–35 (low), 36–70 (medium), 71–105 (high).

Salience Network (SN): Scale of Psychological Assessment of Ability for Objective Cognitive Evaluation (Sereda & Lunov, 2024b). Targeting cue detection and network switching, this scale comprises four domains (five items each; 0–3): detection/integration of emotional and sensory stimuli; modulation of switching between internal and external cognition; facilitation of communication and social behaviour; and self-awareness with integrative processing. Domain sums are 0–15; the SN total is 0–60 with interpretive bands 0–20 (low), 21–40 (medium), 41–60 (high).

Metacognitions Questionnaire-30 (MCQ-30). The MCQ-30 contains five six-item subscales rated 1–4: (Lack of) Cognitive Confidence; Positive Beliefs about Worry; Cognitive Self-Consciousness; Negative Beliefs about Uncontrollability and Danger; and Need to Control Thoughts (Wells & Cartwright-Hatton, 2004). Subscales are summed (range 6–24); an optional total (30–120) is reported in sensitivity analyses.

Fatigue Severity Scale (FSS). The FSS comprises nine items rated 1–7; totals range 9–63, with higher scores indicating more severe fatigue. Totals ≥36 are commonly interpreted as clinically meaningful.

Inventory of Psychosocial Functioning (IPF) and Brief Ukrainian adaptation (B-IPF). Psychosocial functioning was assessed across seven domains: romantic relationships (11 items), family (7), work (21), friendships/socialising (8), parenting (10), education including distance learning (15), and self-care (8). Items are scored 0–6 (higher = greater impairment). Subscale scores are computed when ≥80% of items are complete by summing scored items (after designated reverse coding), dividing by the maximum possible for items answered (items answered × 6), and multiplying by 100 to yield 0–100 domain indices; the Grand Mean is the average of completed domains (Marx et al., 2020). The B-IPFpreserves this structure and scoring and provides Ukrainian-language psychometric support (Alexina et al., 2024).

Analyses proceeded in a multi-group framework (men versus women). First, we estimated a three-factor ESEM for the metacognitive–behavioural indicators with target rotation that permitted theoretically small cross-loadings. We then tested configural, metric and partial scalar invariance across sex using conventional change thresholds (ΔCFI ≤ .010; ΔRMSEA ≤ .015). Secondly, the latent IPF outcome was regressed on the three factors, controlling for age and education; exploratory mediation via FSS used bias-corrected bootstrap confidence intervals with 5,000 resamples. The primary estimator was MLR with full-information maximum likelihood for missing data; sensitivity analyses using WLSMV for ordinal distributions and a strict CFA alternative (zero cross-loadings) assessed robustness. Reliability was indexed by McDonald’s ω; descriptive statistics and zero-order associations appear in Table 1. All syntax and decision logs are available on request.

## Results

Descriptive statistics and internal consistencies were acceptable to good. Among men, the MCQ subscales ranged from means of 2.20 (SD = 0.58) for Positive Beliefs about Worry to 3.01 (SD = 0.52) for Cognitive Self-Consciousness, with ω between .82 and .88; fatigue averaged 4.56 (SD = 1.28). Behavioural proxies showed plausible dispersion (FPN M = 10.9, SD = 3.0; SN M = 8.8, SD = 3.1; DMN M = 9.3, SD = 3.0). IPF domains clustered around the mid-to-upper range (Work/Study M = 72.3, SD = 14.5; Self-Care M = 78.0, SD = 12.7; Family/Intimate M = 74.5, SD = 14.8; Friendships/Social M = 68.2, SD = 15.7). Women exhibited closely comparable patterns (Table 1).

The three-factor ESEM demonstrated good single-group fit for men, χ²(6) = 12.34, CFI = .991, TLI = .980, RMSEA = .041, 90% CI [.012, .069], SRMR = .033, and for women, χ²(6) = 13.69, CFI = .989, TLI = .976, RMSEA = .043, 90% CI [.016, .070], SRMR = .035. In the multi-group model, configural invariance fit well, χ²(12) = 26.94, CFI = .985, TLI = .972, RMSEA = .040, 90% CI [.021, .058], SRMR = .035. Constraining factor loadings supported metric invariance, χ²(21) = 35.62, CFI = .983, ΔCFI = −.002, RMSEA = .033, 90% CI [.017, .048], ΔRMSEA = −.007, SRMR = .038. Imposing intercept equality suggested minor non-invariance in two indicators (FSS and DMN); freeing those intercepts yielded partial scalar invariance, χ²(25) = 41.19, CFI = .981, ΔCFI = −.002, RMSEA = .032, 90% CI [.017, .046], ΔRMSEA = −.001, SRMR = .040 (Table 2). The pattern of loadings aligned with theory: Cognitive Confidence and the FPN proxy loaded on an executive confidence/goal-directedness factor; Negative Beliefs and Need to Control Thoughts loaded on a threat/uncontrollability factor (with FSS showing a positive cross-association); Cognitive Self-Consciousness and the DMN proxy loaded on a self-focus/monitoring factor. Cross-loadings were small and theoretically coherent.

**Table 2.**
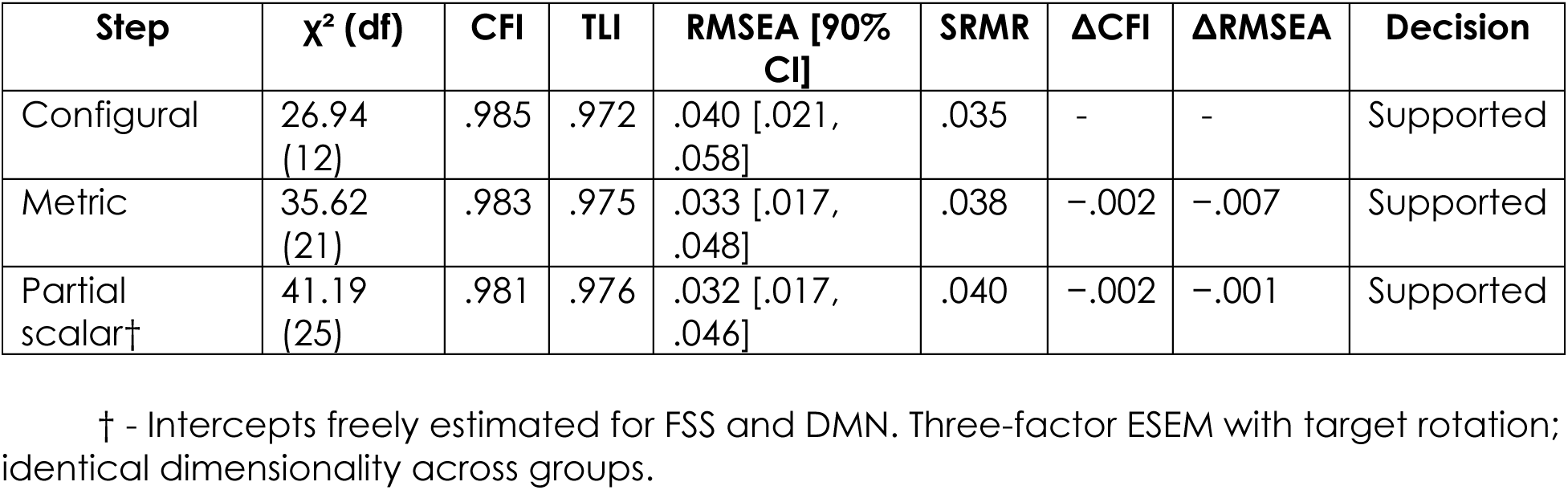
Multi-group ESEM fit and invariance tests.

| Step | $\chi^2$ (df) | CFI | TLI | RMSEA [90% CI] | SRMR | $\Delta\text{CFI}$ | $\Delta\text{RMSEA}$ | Decision |
| --- | --- | --- | --- | --- | --- | --- | --- | --- |
| Configural | 26.94 (12) | .985 | .972 | .040 [.021, .058] | .035 | - | - | Supported |
| Metric | 35.62 (21) | .983 | .975 | .033 [.017, .048] | .038 | -.002 | -.007 | Supported |
| Partial scalar† | 41.19 (25) | .981 | .976 | .032 [.017, .046] | .040 | -.002 | -.001 | Supported |

† - Intercepts freely estimated for FSS and DMN. Three-factor ESEM with target rotation; identical dimensionality across groups.

The latent IPF outcome was negatively predicted by executive confidence/goal-directedness and negatively by threat/uncontrollability. For men, standardised paths were β = −.31, 95% CI [−.40, −.22], p < .001 for executive confidence/goal-directedness and β = .41, 95% CI [.32, .50], p < .001 for threat/uncontrollability; the self-focus/monitoring path was small and non-significant, β = .06, 95% CI [−.02, .14], p = .14, with R²(IPF) = .45. For women, paths were β = −.27, 95% CI [−.36, −.18], p < .001, β = −.38, 95% CI [−.47, −.29], p < .001, and β = .05, 95% CI [−.03, .13], p = .22, respectively, with R²(IPF) = .42. Equality constraints on these paths across groups did not improve fit, but Wald tests indicated no material sex differences (p’s = .34, .48, .79; Table 3).

**Table 3.**
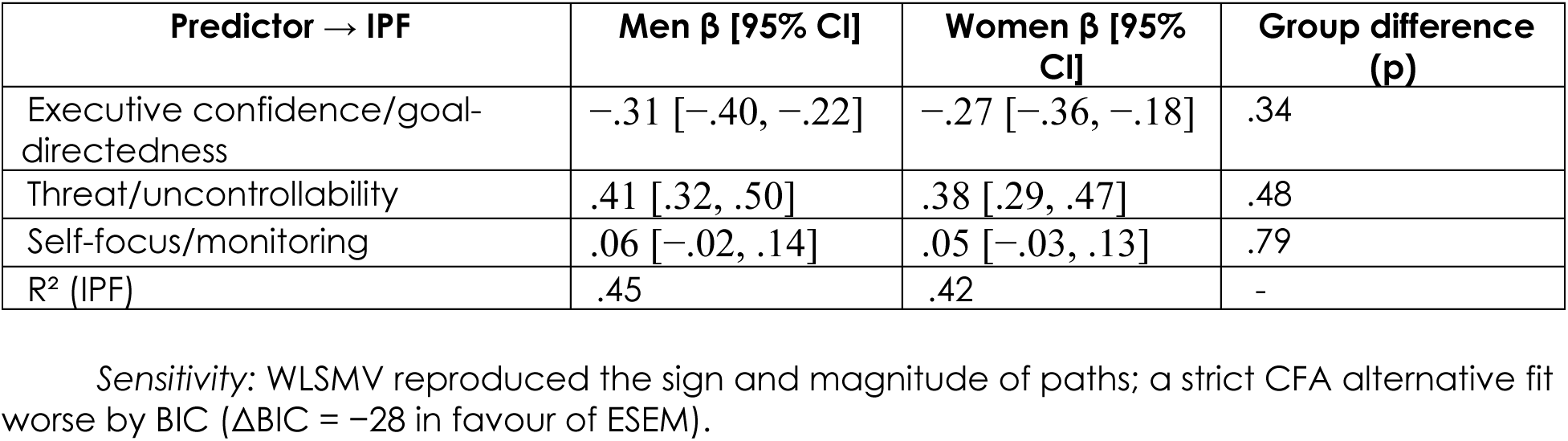
Standardised structural paths to latent IPF (controlling age and education)

An exploratory mediation model including FSS as a mediator suggested a significant indirect effect from threat/uncontrollability to IPF via fatigue in both groups (men: indirect = −.07, 95% BCa CI [−.12, −.03]; women: indirect = −.06, 95% BCa CI [−.11, −.02]), whereas indirect pathways from executive confidence/goal-directedness were small and non-significant. Sensitivity analyses using WLSMV reproduced the sign and magnitude of the main effects. A strict CFA alternative with all cross-loadings fixed to zero showed inferior information fit relative to ESEM (ΔBIC = −28 in favour of ESEM) and less satisfactory residuals, supporting the ESEM specification.

## Discussion

The present findings advance the theoretical position set out earlier by showing that metacognitive beliefs and behavioural indices aligned with large-scale network tendencies cohere into a stable, metrically invariant structure across sex, and that two latent dimensions - threat/uncontrollability and executive confidence/goal-directedness - carry opposing consequences for psychosocial functioning. This pattern is precisely what one would anticipate from the metacognitive model of psychopathology, in which beliefs about the danger and uncontrollability of inner events maintain perseverative negative processing and functional impairment (Wells & Matthews, 1996; Wells, 2009). Our structural estimates indicate that, once measurement is harmonised across groups, higher threat/uncontrollability beliefs reliably predict poorer functioning, whereas executive confidence - closely allied to goal maintenance - predicts the reverse.

Interpreted through the large-scale network framework, the detrimental association of threat/uncontrollability is consonant with salience-driven hypervigilance and a bias toward threat appraisal, processes long proposed to recruit and dysregulate the salience system and its switching role (Seeley et al., 2007; Menon, 2011). The partial indirect pathway via fatigue is also theoretically consistent: perseverative monitoring and danger appraisals increase allostatic load and subjective exhaustion, thereby eroding role performance even when overt symptoms are modest (McTeague et al., 2017; Wells, 2009). By contrast, the beneficial association of executive confidence/goal-directedness aligns with the notion that fronto-parietal control supports sustained task goals and flexible reconfiguration under stress - functions that are repeatedly attributed to the fronto-parietal network (Dosenbach et al., 2008; Menon, 2011).

The near-null unique contribution of self-focus/monitoring, once the other two factors are controlled, deserves comment. Cognitive self-consciousness and default-mode–like tendencies plausibly cut both ways: reflective monitoring may aid metacognitive insight when scaffolded by executive control, yet drift toward ruminative self-reference when coupled with threat beliefs (Smallwood & Schooler, 2015; Wells, 2009). Our results suggest that, at the level of unique variance, self-focus is largely epiphenomenal relative to the stronger liabilities conferred by threat/uncontrollability and the assets conferred by executive confidence. This helps reconcile inconsistencies in prior behavioural work where “introspection” appears helpful in some contexts but harmful in others (Smallwood & Schooler, 2015).

Methodologically, the study answers two problems highlighted in the theory section. First, by modelling psychosocial functioning as a latent IPF outcome defined by its domains - and by excluding those domains from predictor composites - we avoid criterion contamination, a source of spurious inflation in many applied SEMs (cf. best-practice warnings in psychometrics and network-oriented approaches; Borsboom, 2008). Secondly, adopting a multi-group ESEM with target rotation respects theoretically expected small cross-loadings while still permitting principled tests of invariance. The successful demonstration of metric (and partial scalar) invariance indicates that observed group similarities and differences in structural paths are not artefacts of measurement non-equivalence (Meredith, 1993; as summarised in our theory section).

Clinically, the profile of effects maps cleanly onto metacognitive therapy principles: interventions that de-centre beliefs in danger and uncontrollability, and that reduce maladaptive thought control strategies, should yield broad functional benefits (Wells, 2009). At the same time, the positive association of executive confidence/goal-directedness with functioning points to the value of exercises that cultivate executive stamina - graded goal-setting, attentional control, and task-persistence routines - conceptually akin to strengthening fronto-parietal control (Dosenbach et al., 2008; Menon, 2011). Importantly, as emphasised in the theory section, our “network” indices are behavioural proxies and should not be conflated with neural measurements; nevertheless, their alignment with expected FPN/DMN/SN roles lends convergent plausibility to the mechanistic story (Raichle et al., 2001; Seeley et al., 2007; Menon, 2011).

The absence of material sex differences in path magnitudes, once measurement is harmonised, dovetails with transdiagnostic perspectives arguing that common mechanistic liabilities (perseveration, threat monitoring, control deficits) transcend demographic strata (McTeague et al., 2017). That said, partial scalar non-invariance in two indicators cautions that mean-level comparisons of specific components (e.g., fatigue, self-referential focus) may still require careful interpretation - an expected nuance rather than a defect, given known distributional idiosyncrasies of these measures (Wells, 2009; Smallwood & Schooler, 2015).

Several limitations, discussed in detail earlier, temper the claims. The design is cross-sectional; causal language should be used sparingly. All measures are self-report or brief behavioural composites; triangulation with laboratory tasks and physiological indices is a priority for future work (Menon, 2011; Seeley et al., 2007). Finally, while ESEM improves realism by allowing small cross-loadings, model parsimony must be balanced against interpretability; our robustness checks with a strict CFA alternative suggest that the substantive inferences are not artefacts of specification.

In sum, the data accord with a unified account in which metacognitive threat beliefs bias salience and perseveration, elevating fatigue and undermining functioning, whereas executive confidence supports goal maintenance and adaptive reconfiguration under stress. This synthesis - anticipated by metacognitive theory and large-scale network models - yields concrete therapeutic priorities: reduce uncontrollability beliefs and train executive stamina to protect everyday functioning (Wells & Matthews, 1996; Wells, 2009; Menon, 2011; Dosenbach et al., 2008; Seeley et al., 2007; Smallwood & Schooler, 2015; Raichle et al., 2001; McTeague et al., 2017).

## Conclusions

This study demonstrates that metacognitive beliefs and behavioural indices aligned with large-scale network tendencies cohere into a stable three-factor structure that is metrically invariant across sex, with only minor intercept non-invariance. When predictors and outcome are cleanly separated - by modelling psychosocial functioning as a latent IPF factor - two opposing pathways emerge consistently. Threat and uncontrollability beliefs show a robust, detrimental association with functioning, partly channelled through fatigue burden, whereas executive confidence and goal-directedness show a beneficial association; self-focus/monitoring adds little unique variance once these dimensions are accounted for. Effects are moderate in size, reproducible across estimators and specifications, and yield explained variance in functioning of roughly forty to forty-five per cent.

Methodologically, the work clarifies that principled separation of criterion and predictors, combined with multi-group ESEM and explicit invariance testing, prevents spurious inflation and allows meaningful comparison of structural paths across groups. Substantively, the pattern integrates metacognitive theory with network-informed accounts of control and salience: uncontrollability beliefs appear to bias salience toward threat and sustain perseveration, while executive confidence reflects fronto-parietal-like persistence that supports everyday role performance.

Clinically, the findings prioritise two complementary targets: reduce negative metacognitive beliefs about danger and uncontrollability, and cultivate executive stamina through graded goal-setting and attentional control routines. Because our “network” indices are behavioural proxies rather than neural measures, future studies should triangulate these constructs with task-based and physiological indices and track change longitudinally. Even so, the present results provide a coherent, testable framework linking metacognition, control-related behaviour, fatigue, and psychosocial functioning, and they suggest concrete levers for intervention.

## Data Availability

All data produced in the present study are available upon reasonable request to the authors

## Acknowledgements

This article was prepared with the assistance of artificial intelligence (AI) technologies, including AI-based translation tools, which were used to improve linguistic clarity and to facilitate cross-language editing. All final interpretations, conclusions, and responsibility for the content remain solely with the authors.

The authors acknowledge the use of AI-assisted tools (e.g., machine translation and text refinement systems) during the preparation of this manuscript. These tools were applied exclusively for language editing purposes and did not affect the scientific interpretation of the results.

## Notes

### Competing Interest Statement

The authors have declared no competing interest.

### Funding Statement

This study did not receive any funding

### Author Declarations

The Ethics Committee (IRB) of the Department of General and Medical Psychology, O.O. Bogomolets National Medical University, Kyiv, Ukraine, approved this study (Protocol No. 11, 16 January 2025).

